# Association between Overactive Bladder and Lipid Accumulation Products and Visceral Adiposity Index: Results from NHANES 2005-2018

**DOI:** 10.1101/2025.03.05.25323429

**Authors:** Wenhao Wang, Xiaolin Xu

## Abstract

**Background:** Although obesity is a recognized risk factor contributing to the onset and progression of overactive bladder (OAB), the existing evidence linking lipid accumulation products (LAPs) and the visceral adiposity index (VAI) to OAB remains scarce and subject to debate. Hence, this study was conducted to evaluate the associations between VAI, LAP, and the occurrence of OAB.

**Methods:** The information utilized in this research was sourced from the National Health and Nutrition Examination Survey (NHANES), spanning the years 2005 to 2018. The majority of the data pertaining to OAB relied on self-administered questionnaires. To assess the relationships between VAI, LAP, and OAB, we employed multivariate logistic regression models, trend analysis, and subgroup evaluations.

**Results:** The study encompassed a total of 70,190 participants, with 22,928 individuals diagnosed with OAB and 5,776 serving as controls. After accounting for potential confounding factors, a statistically significant positive relationship was noted between both the visceral adiposity index (VAI) and the lipid accumulation product (LAP), as well as the occurrence of overactive bladder (OAB). Respectively, individuals in the highest quartiles of LAP and VAI demonstrated a 56% (OR = 1.555, 95% CI: 1.376 to 1.758) and 22% (OR = 1.225, 95% CI: 1.084 to 1.384) increased probability of OAB when compared to those in the lowest quartile. Additional subgroup analyses revealed that the observed associations were particularly evident in participants under the age of 60 and among women.

**Conclusion:** This study’s findings suggest that an increase in both the visceral adiposity index (VAI) and lipid accumulation product (LAP) is associated with a greater occurrence of overactive bladder (OAB), hinting at their possible use as predictive indicators for OAB.

## Introduction

Primarily characterized by urinary urgency, overactive bladder (OAB) is a clinical syndrome that manifests in the absence of urinary tract infections or any other underlying medical conditions. It is often associated with increased frequency and nocturia, with or without urgency urinary incontinence[1][2]. According to epidemiological research, the incidence of OAB stands at 10.8% among males and 12.8% among females.[3]. Furthermore, the incidence of OAB escalates with advancing age. As the global population expands and ages, the socioeconomic impact of OAB is progressively intensifying.[4]. The underlying pathophysiological mechanisms of OAB are still not fully elucidated. However, current research indicates that a multitude of factors, such as neuromodulation of the bladder, muscular tissue involvement, central nervous system regulation, inflammatory processes, and metabolic disturbances, may collectively play a role in the development of this condition.[5]. Consequently, identifying novel and more precise biomarkers for the early diagnosis and prevention of OAB is highly important.

Numerous studies have emphasized the crucial part obesity plays in the onset and advancement of OAB[6]. A key characteristic of obesity is the accumulation of visceral adipose tissue. Nonetheless, conventional metrics like body mass index (BMI) and waist circumference (WC) offer only a rough estimation of obesity and lack the capacity to differentiate between subcutaneous and visceral fat deposits[7]. The lipid accumulation product (LAP) incorporates measures of waist circumference and triglyceride levels, serving as an indicator of adipose tissue accumulation and metabolic impairment[8]. Waist circumference, BMI, high-density lipoprotein (HDL) cholesterol levels, and triglyceride concentrations are all taken into consideration by the visceral adiposity index (VAI), which evaluates the distribution and functioning of visceral fat[9]. These two indices act as reliable markers of visceral fat buildup and metabolic disorders. They quantify obesity and hint at visceral fat’s impact on bladder function.[10][11]. In specific clinical scenarios, their predictive capability may even exceed that of traditional measurements like BMI[12]. Previous studies have also established associations between LAP, VAI, and other diseases[13][14].

The foundation of this study is the National Health and Nutrition Examination Survey (NHANES) database, a comprehensive clinical dataset meticulously maintained by the National Center for Health Statistics (NCHS). Leveraging data from the NHANES database encompassing the years 2005 to 2018, this research endeavors to evaluate the potential association between LAP, VAI, and OAB, ultimately aiming to facilitate early diagnosis and prevention of OAB.

## Methods

### Study population

The data for this analysis originated from the National Health and Nutrition Examination Survey (NHANES), conducted between 2005 and 2018. NHANES is a research endeavor administered by the Centers for Disease Control and Prevention (CDC), intended to gather exhaustive information on the health status, dietary habits, and lifestyles of the U.S. populace. It employs a sophisticated, multi-stage stratified sampling technique to collect these data. The protocols employed in NHANES were cleared by the Review Board of the National Center for Health Statistics (NCHS). All participants provided their informed consent. The NHANES database encompasses five key components: demographic information, screening results, dietary records, laboratory test outcomes, and questionnaire responses [15]. Additional information on the NHANES database is accessible at http://www.cdc.gov/nhanes.

In total, 70,190 participants were incorporated into the NHANES dataset spanning from 2005 to 2018. Figure 1 depicts the exclusion criteria, which include: (1) participants with absent OAB data; (2) those lacking information on VAI and LAP; and (3) individuals missing covariate data or below the age of 20. Consequently, a final count of 28,704 samples was retained for the definitive analysis. The procedure for data selection is illustrated in Figure 1.

**Fig. 1.**
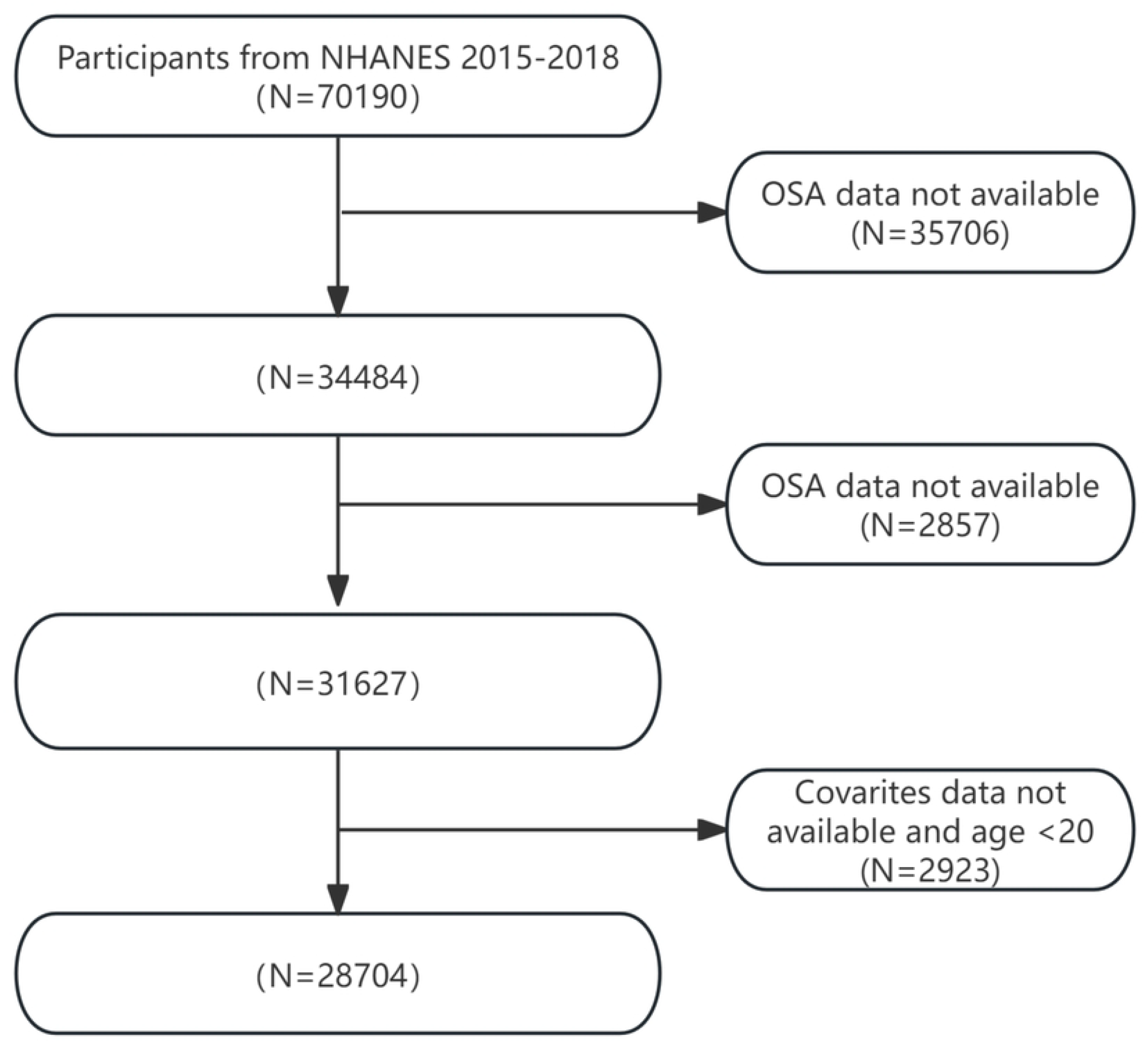
Flow chart of patient screening.

### Definition of Exposure Variables

In this study, VAI and LAP were designated as the exposure variables. NHANES involved the measurement of triglyceride (TG) and high-density lipoprotein cholesterol (HDL-C) levels using fasting blood samples, which were collected by healthcare professionals at the Mobile Examination Centers (MECs).At the Mobile Examination Centers (MECs), waist circumference (WC) was assessed by skilled technicians using a tape measure, positioned at the midpoint of the axillary line just above the iliac crest during the end of a normal exhalation, with a precision of 0.1 cm. The calculations for LAP and VAI were gender-specific and based on the following formulas, where TG and HDL were measured in mmol/L, WC in centimeters, and BMI in kilograms per square meter[16].

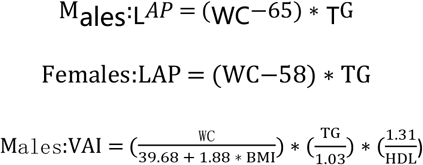

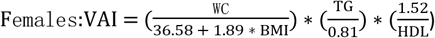

### Definition of OAB

Overactive bladder (OAB) is a condition primarily characterized by urinary frequency, urgency urinary incontinence (UUI), and nocturia. In this study, standardized questionnaires were administered during face-to-face interviews conducted by trained personnel to collect the necessary data.The severity of UUI was determined by two questions: “In the past 12 months, have you experienced urine leakage or loss of bladder control due to urinary urgency or pressure, and been unable to reach the toilet in time?” and “How frequently does this occur?” Nocturia was assessed on the basis of the following question: “In the past 30 days, how many times did you typically wake up at night to urinate, from the time you went to bed until you got up in the morning?” Ultimately, the severity of OAB for each participant was assessed using the Overactive Bladder Symptom Score (OABSS), where a total score of three or higher was deemed indicative of the presence of OAB[17].

### Definition of Covariates

Drawing from the existing literature, the following variables were identified as covariates: gender, age, ethnicity, educational attainment, marital situation, income-to-poverty ratio, smoking habits, alcohol intake, and the occurrence of hypertension or diabetes. Ethnicity was grouped into non-Hispanic White, non-Hispanic Black, Mexican American, other Hispanic, and other racial categories. Marital situation was categorized as either living alone or being married/in a cohabiting relationship. The educational level is classified into three categories: below high school, high school, or above high school. The poverty-to-income ratios (PIRs) were categorized as <1.5, 1.5–3.5, and >3.5. Smoking and alcohol consumption were determined on the basis of the following questions: “Have you ever smoked at least 100 cigarettes in your lifetime?” and “Do you consume at least 12 alcoholic beverages per year?”[18].

### Statistical Analysis

In this study, the NHANES-recommended sample weights were utilized for analysis. Continuous variables were presented as the mean, accompanied by their standard deviations, whereas categorical variables were denoted as percentages. The LAP and VAI data were stratified into quartiles, with the lowest quartile serving as the baseline comparison group. To explore the associations between LAP, VAI, and OAB, multivariable logistic regression models were employed to estimate odds ratios (ORs) and their corresponding 95% confidence intervals (CIs) for each variable. Model 1 served as the unadjusted baseline, without considering any confounding variables. Model 2 incorporated adjustments for sex, age, race, educational level, marital status, and poverty-income ratio (PIR). Model 3 extended these adjustments by further accounting for smoking history, alcohol use, hypertension, and diabetes, in addition to the variables included in Model 2. Subgroup analyses were carried out based on sex, age, race, educational attainment, smoking history, alcohol intake, and diabetes status. All statistical assessments were executed using EmpowerStats version 4.2 software, with all tests conducted on a two-tailed basis. Statistical significance was established at a p value of less than 0.05.

## Results

### Baseline characteristics of participants

Table 1 Baseline characteristics of participants in the NHANES 2005–2018

LAP: Lipid accumulation product, VAI:Visceral adiposity Index, NHANES: National Health and Nutrition Examination Survey *p < 0.05.

Table 1 summarizes the baseline characteristics of the participants. On the basis of the data analysis of 28,704 participants who met the inclusion and exclusion criteria, 5776 (20.12%) were diagnosed with overactive bladder syndrome (OAB), and 22958 (79.88%) did not have OAB. Among these participants, 62.88% were women, with a mean age of 47.21 years. Compared with participants without OAB, those with OAB were more likely to be female, older, have a larger waist circumference, live alone, have a lower educational level, be diagnosed with diabetes, and have higher LAP and VAI values. (all p < 0.05).

### Associations between LAP, VAI, and OAB

Table 2 Multivariate logistic regression analysis of LAP and VAI with overactive bladder

Abbreviations: OAB: Overactive bladder.; VAI: Visceral adiposity index; LAP: Lipid accumulation product; NHANES: National Health and Nutrition Examination Survey; OR: odds ratio: CI: confidence interval

The results of the multivariable linear regression analysis are shown in Table 2. The results indicate a notable link between both LAP and VAI and the likelihood of developing OAB. We analyzed three different models and categorized the LAP and VAI indices into four quartiles. According to the unadjusted model (Model 1), higher LAP and VAI values were associated with an increased incidence of OAB. The odds ratios (ORs) for the highest quartile of LAP (OR = 2.407, 95% CI: 2.150, 2.695) and the OR for the highest quartile of VAI (OR = 1.726, 95% CI: 1.539, 1.936) were both significantly greater than those for the lowest quartile, with increases of 141% and 72%, respectively. After fully adjusting for all confounding factors (Model 3), the significant associations between LAP, VAI, and OAB persisted.The OR for the highest quartile of LAP (OR = 1.555, 95% CI: 1.376, 1.758) was 56% higher than that for the lowest quartile, with a trend p-value of < 0.0001. The OR for the highest quartile of the VAI (OR = 1.225, 95% CI: 1.084, 1.384) was 22% higher than that for the lowest quartile, with a trend p value of < 0.0001.

Table 3 Threshold effect analysis of LAP and VAI on OAB in the NHANES 2005–2018

Notes: Age, race, sex, level of education, marital status, poverty income ratio, alcohol consumption, smoking status, hypertension statues and diabetes were adjusted

The solid red line signifies the fitted smooth curve that represents the relationship between the variables, and the blue bands denote the 95% confidence interval associated with this fitting.

To further visualize the relationships among the LAP, VAI, and OAB, we performed smooth curve fitting with Model 3. Figure 2 illustrates the nonlinear associations among the LAP, VAI, and OAB. A threshold effect analysis was subsequently conducted to clarify the associations (Table 3).The turning point for LAP was identified at 86.234, with a likelihood ratio < 0.001, indicating that when LAP levels drop below 86.23, each unit increase in LAP is associated with a 0.6% increase in OAB risk. When LAP levels exceeded 86.234, the correlation between LAP and OAB risk diminished, suggesting that further increases in LAP did not lead to a statistically significant increase in OAB risk.The results for the VAI exhibited a similar pattern, with a turning point at 1.49 (likelihood ratio 0.025), suggesting that the VAI had different effects on OAB risk below and above this threshold.

**Fig. 2.**
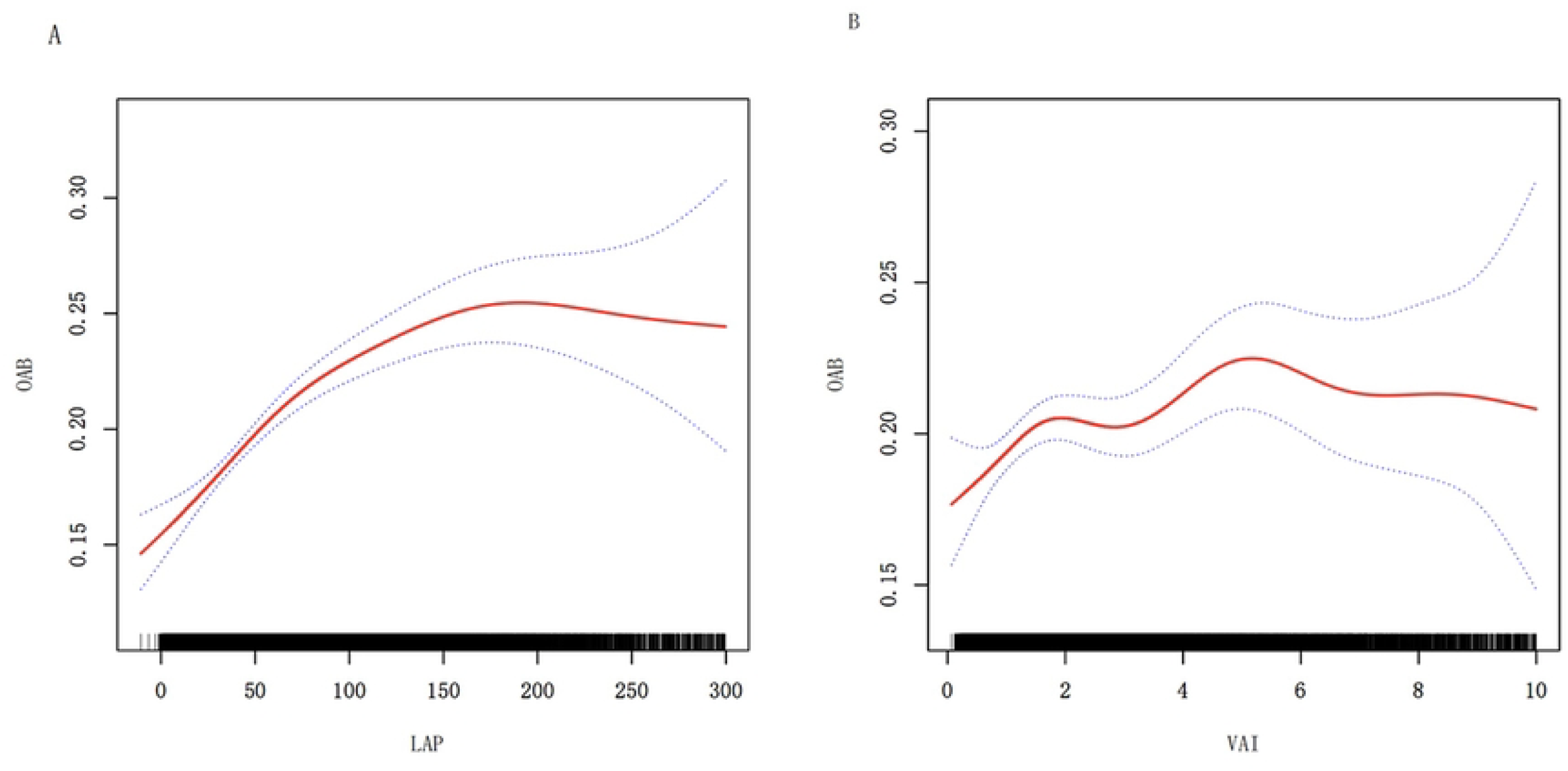
Nonlinear relationships between LAP and VAI and OAB.

### Subgroup analysis

Table 4 Subgroup analyses

Abbreviations: OR: odds ratio; CI: confidence interval

Notes: Age, race, sex, level of education, marital status, poverty income ratio, alcohol consumption, smoking status, hypertension statues and diabetes were adjusted. The strata variable was not included in the model when stratifying by itself

To evaluate the stability of the associations among the VAI, LAP, and OAB, additional subgroup analyses were performed. Subgroup analyses were stratified by sex, age < 60 years, education level, smoking status, alcohol consumption, and the presence of diabetes. The results of these subgroup analyses, adjusted for all confounding factors, are presented in Table 4. The associations between LAP, VAI, and OAB were consistent across all subgroups.

It is worth noting that there were notable interactions detected among LAP, VAI, as well as gender and age, with statistical significance at p < 0.05. A statistically significant positive correlation was found in women, but this association was not observed in men, indicating that increased visceral fat accumulation may increase the likelihood of OAB in women. Additionally, significant interactions between LAP, VAI, and age < 60 years were found, whereas the results became nonsignificant in participants aged ≥ 60 years, suggesting that higher levels of LAP and VAI may increase the risk of OAB in individuals aged < 60 years.

## Discussion

This study explored the relationship between two visceral adiposity indices—Lipid Accumulation Product (LAP) and Visceral Adiposity Index (VAI)—and overactive bladder (OAB). The sample included 28,704 participants. The findings revealed a significant positive correlation between both LAP and VAI with OAB. Additionally, a nonlinear relationship was observed, with specific threshold points for each index. These results suggest that within certain value ranges, LAP and VAI may have clinical relevance in reducing the risk of OAB.

The relationship between OAB and obesity is complex and interdependent[19]. Numerous studies have established a connection between obesity and the development of OAB. For instance, a meta-analysis found that women with a body fat percentage above 32% have a 95% increased risk of developing OAB compared to those with a body fat percentage below this threshold[20]. Further research indicates that an increase in LAP is linked to an elevated risk of stress urinary incontinence[21]. Additionally, the accumulation of abdominal fat, particularly in the context of obesity, is a significant risk factor for both the onset and progression of OAB[22]. Increased abdominal pressure associated with obesity can compress the bladder, reducing its capacity and ability to store urine. This compression may impair bladder function and contribute to the development of OAB[23]. Obesity is also linked to a chronic state of low-grade systemic inflammation. Increased visceral fat tissue releases various pro-inflammatory cytokines, which can disrupt the neuromuscular function of the bladder. This disruption may, in turn, raise the risk of developing OAB[24]. In addition to physical compression and inflammatory factors, obesity is often associated with metabolic syndrome. Conditions such as insulin resistance in the bladder mucosa, hyperglycemia, and hormonal imbalances (including estrogen and leptin) can alter bladder blood flow and neural regulation. These changes may contribute to the onset of OAB[25][26][27].

Compared to traditional urodynamic evaluations, the LAP and VAI provide simpler, more convenient, and cost-effective methods for diagnosing OAB[28][29]. Research has demonstrated a lack of substantial correlation between either waist circumference (WC) or body mass index (BMI) and OAB symptoms[30]. Hence, when compared to conventional approaches like BMI and WC, the LAP and VAI exhibit enhanced precision and predictive power[31]. Recent research indicates that both the LAP and VAI can function as indicators of insulin resistance and metabolic disturbances[32][33][34]. Additionally, studies have demonstrated that LAP is linked to insulin resistance, lipid peroxidation, and systemic inflammatory responses in individuals with type 2 diabetes, all of which constitute risk factors for the development of OAB[35]. Our research has also established a positive relationship between LAP, VAI, and OAB, hinting at their potential role as diagnostic and preventive benchmarks in clinical practice for OAB. Furthermore, prior studies have suggested that abdominal obesity correlates with a heightened vulnerability to moderate to severe lower urinary tract symptoms among adults[36]. A study with a cross-sectional design, encompassing 27,309 participants, has uncovered the potential of the VAI to serve as a predictive marker for insulin resistance[37]. Collectively, these studies establish a groundwork for elucidating the connection between visceral fat accumulation and OAB. Our research utilizes the LAP and VAI, indices indicative of visceral fat accumulation, as diagnostic tools for assessing OAB symptoms and their severity.

Subgroup analyses conducted within this study revealed that elevated levels of LAP and VAI were linked to a heightened risk of OAB among participants younger than 60 years and in females. However, this correlation seemed to diminish among individuals aged 60 and above, as well as in males. This observation could potentially be attributed to age-induced alterations in physiological and metabolic processes. In older adults, the presence of comorbidities such as hypertension, diabetes, and cardiovascular disorders may diminish the strength of the relationships between LAP, VAI, and OAB[38]. Tung Wai Auyeung and colleagues confirmed that older adults may exhibit some resistance to the effects of overweight and obesity, with even mild obesity or overweight potentially offering some protection[39]. Moreover, women typically possess a higher fat mass relative to their body weight compared to men, and gender-specific differences exist in the association between obesity and the incidence of OAB[40][41].

### Study strengths and limitations

Our study boasts several notable strengths. Leveraging the NHANES database from the United States, a cross-sectional approach was employed, benefiting from its highly dependable data owing to stringent quality assurance and standardized methodologies for data acquisition. Furthermore, the utilization of LAP and VAI as indicators of visceral fat distribution offers simplicity, cost-effectiveness, and enhanced precision compared to conventional metrics like WC and BMI, which may be prone to inaccuracies. These indices possess the potential to aid physicians in tailoring more individualized treatment strategies for patients, based on their specific LAP and VAI scores.

Although our study has identified a possible correlation between LAP, VAI, and OAB, it is important to acknowledge several constraints. Primarily, the cross-sectional design of the study prevents the establishment of a definitive causal link between LAP, VAI, and OAB. Secondly, the diagnosis of OAB was largely reliant on self-reported questionnaire responses, which could potentially introduce bias stemming from recall inaccuracies. Lastly, and crucially, the study’s survey data is predominantly from the U.S. population, necessitating further validation to ascertain the generalizability of these findings to populations in diverse geographical regions.

## Conclusion

In summary, using NHANES data, our study has demonstrated a notable correlation between increased LAP and VAI values and the likelihood of developing OAB, suggesting that these indicators could function as valuable predictive biomarkers for OAB. For those with elevated LAP and VAI scores, a holistic approach encompassing dietary adjustments and exercise interventions is advisable to postpone and manage OAB, or specific strategies should be undertaken to forestall and enhance outcomes associated with OAB.

## Acknowledgements

We acknowledge with gratitude the contributions made by the participants and survey staff of the National Health and Nutrition Examination Survey (NHANES).

## Author contributions

Wenhao Wang conceived the study design, conducted the data analysis, and drafted the manuscript. Xiaolin Xu contributed ideas for the study and refined the final version of the manuscript. All authors reviewed and endorsed the final manuscript.

## Data availability

The datasets that were produced and examined in this research can be accessed on the NHANES website, located at: https://www.cdc.gov/nchs/nhanes/index.htm.

## Declarations

## Ethical statement

The NHANES website officially provides access to all data analyzed in this experiment, with all participants having completed the necessary informed consent forms and having undergone the ethical review process, as stipulated by policy.

## Human Ethics and Consent to Participate Statement

Not applicable

## Clinical Trial Number

Not applicable

## Contributor Information

Wenhao Wang, Xiaolin Xu

